# Prevalence of deleterious cardiomyopathy variants in early-onset atrial fibrillation

**DOI:** 10.1101/2025.03.12.25323872

**Authors:** Quim Bech Vilaseca, Oliver Bundgaard Vad, Christian Paludan-Müller, Laura Andreasen, Morten Salling Olesen, Jesper Hastrup Svendsen, Pia Rengtved Lundegaard

## Abstract

**Background:** Atrial fibrillation (AF) is a common cardiac arrhythmia associated with an increased risk of stroke, heart failure, and death. Recent studies suggests that individuals with early onset of AF could be at increased risk of developing heart failure and dilated cardiomyopathy. This study aimed to identifying genetic variants in a broad panel of cardiomyopathy genes among early-onset AF individuals.

**Methods:** We conducted targeted genetic sequencing of 29 cardiomyopathy-associated genes in 478 individuals with AF onset below 45 years of age from a Danish cohort. Additionally, we analyzed whole exome sequencing data in 374,289 individuals from the UK Biobank, including 29,108 individuals with AF. The cohort was stratified by age at AF diagnosis, and individuals with pre-existing cardiomyopathy were excluded. We focused on rare, truncating variants predicted to lead to loss of function, and potentially deleterious missense variants in the UK Biobank.

**Results:** In the Danish cohort, 42 (8.8%) individuals with early-onset AF had truncating genetic variants in known cardiomyopathy genes. The UK Biobank analysis showed an inverse dose-response-like relationship between age of AF onset and prevalence of truncating variants, ranging from 3.8% in the AF onset <45 years group to 1.4% in the group without AF diagnosis. The prevalence of rare missense variants showed a similar pattern.

**Conclusions:** We identified a high prevalence of deleterious variants in cardiomyopathy-associated genes among individuals with early-onset AF. This supports recent guideline suggestions and indicates that genetic testing and surveillance for cardiomyopathy could be relevant in selected individuals with an early AF diagnosis.

## Introduction

With a lifetime risk of up to 30%(1), atrial fibrillation (AF) is the most common cardiac arrhythmia, and represents a serious burden for patients and healthcare systems(2). Among individuals with AF, one in five suffers a stroke, and two in five develop heart failure (HF)(1), leading to increased risk of morbidity and mortality.

While AF has been extensively studied, its association with subsequent cardiovascular diseases and mortality across age groups is not entirely understood. A recent study found that the risk of cardiomyopathy and HF was highest among those with AF diagnosis early in life(3). Moreover, an earlier onset of AF was associated with a considerable decrease in life expectancy compared with a later onset of the disease(3). It has been suggested to be due to a shared genetic architecture of AF and cardiomyopathy(3). Atrial cardiomyopathy has been suggested as both an important component in AF pathogenesis, and as a potential precursor to more severe, ventricular cardiomyopathy phenotypes(4,5).

In a cohort of 1,293 individuals with AF before age 65, Yoneda et al. reported an increased prevalence of potentially pathogenic genetic variants in genes associated with arrhythmia syndromes, cardiac structure, and cardiomyopathy(6). Individuals who were carriers of such variants also exhibited an increased mortality compared to non-carriers(7). In line with these findings, the newest AF guidelines from the American College of Cardiology and the American Heart Association now recommend that genetic testing could be considered in early-onset AF patients without obvious risk factors(8). However, the prevalence of cardiomyopathy-associated genetic variants in early-onset AF in large population-based studies has not been thoroughly explored.

This study included next-generation sequencing data from Danish AF patients diagnosed before the age of 45 without known cardiomyopathy and HF, and whole exome sequencing data from 374,289 individuals from the UK Biobank. Focusing on pathogenic variants in well-established cardiomyopathy genes, this study aimed to provide new insights into the burden of rare variants in cardiomyopathy-associated genes according to age of AF onset.

## Methods

The study followed the *Strengthening the Reporting of Observational Studies in Epidemiology* (STROBE) reporting guideline. All participants gave written informed consent. The study of the Danish cohort was approved by the scientific ethics committee of the Capital Region of Denmark (protocol number: H-20048862). The UK Biobank was accessed under application number 43247. The UK Biobank received ethical approval from the North West Multi-Centre Research Ethics Committee. A detailed methods section is available in the **Supplemental Material**.

## Results

This study included 478 Danish individuals and 374,289 participants from the UK Biobank. Baseline characteristics of the cohorts are summarized in **Table 1**. In the Danish cohort, all individuals had an AF diagnosis at time of inclusion, with a median (1^st^-3^rd^ quartile) age at AF diagnosis of 28 (23-34) years. Among them, 85 participants (17.8%) were female. In contrast, the UK Biobank included 29,108 individuals diagnosed with AF and 345,181 participants without an AF diagnosis, who served as controls. The median (1^st^-3^rd^ quartile) age at enrollment in the UK Biobank was 58 (51-63) years, and 201,335 participants (53.8%) were female.

**Table 1.** Baseline characteristics at inclusion in the Danish and UK Biobank cohorts.

|  | Danish cohort<br>(N = 478) | UK Biobank<br>(N = 374,289) |  |  |  |  |
| --- | --- | --- | --- | --- | --- | --- |
|  | AF onset <45<br>years | No AF<br>(reference) | AF onset <45<br>years | AF onset 45-54<br>years | AF onset 55-64<br>years | AF onset >65<br>years |
| **N** | 478 | 345,181 | 684 | 2,464 | 7,775 | 18,185 |
| **Sex** |  |  |  |  |  |  |
| Female, n (%) | 85 (17.8) | 190,832 (55.3) | 207 (30.3) | 715 (29.0) | 2,534 (32.6) | 7,047 (38.8) |
| Male, n (%) | 393 (82.2) | 154,349 (44.7) | 477 (69.7) | 1,749 (71.0) | 5,241 (67.4) | 11,138 (61.2) |
| **Age at AF diagnosis, years,**<br>**median (1<sup>st</sup>-3<sup>rd</sup> quartile)** | 28 (23, 34) | No AF | 40 (36, 43) | 51 (49, 53) | 61 (58, 63) | 71 (68, 75) |
| **BMI, kg/m<sup>2</sup>, median (1<sup>st</sup>-3<sup>rd</sup>**<br>**quartile)** | 26.0 (23.5, 29.0) | 26.6 (24.1, 29.7) | 27.6 (24.8, 31.2) | 28.2 (25.3, 31.9) | 28.3 (25.4, 32.1) | 28.1 (25.3, 31.5) |
| **Blood pressure** |  |  |  |  |  |  |
| Systolic, mmHg, median<br>(1<sup>st</sup>- 3<sup>rd</sup> quartile) | 128 (120, 135) | 138 (126, 152) | 133 (122, 146) | 137 (124, 150) | 140 (127, 153) | 146 (134, 160) |
| Diastolic, mmHg, median<br>(1<sup>st</sup>-3<sup>rd</sup> quartile) | 79 (71, 83) | 82 (75, 89) | 81 (75, 89) | 83 (75, 91) | 83 (75, 90.8) | 82 (75, 90) |
| **Hypertension, n (%)** | 30 (6.3) | 366 (53.5) | 1,488 (60.4) | 5,272 (67.8) | 13,512 (74.3) | 124,154 (36.0) |
| **Diabetes, n (%)** | 6 (1.3) | 27,428 (7.9) | 115 (16.8) | 503 (20.4) | 1,650 (21.2) | 4,024 (22.12) |

AF, Atrial Fibrillation; BMI, Body Mass Index

### High prevalence of truncating variants in the Danish cohort

Among 478 Danish individuals diagnosed with AF before age 45 and no known cardiovascular comorbidities, 42 individuals (8.8%) were carriers of 38 rare truncating variants in well-established cardiomyopathy-related genes (**Figure 1a)**. This cohort exhibited genetic variants in nine of the 29 selected genes.

**Figure 1.**
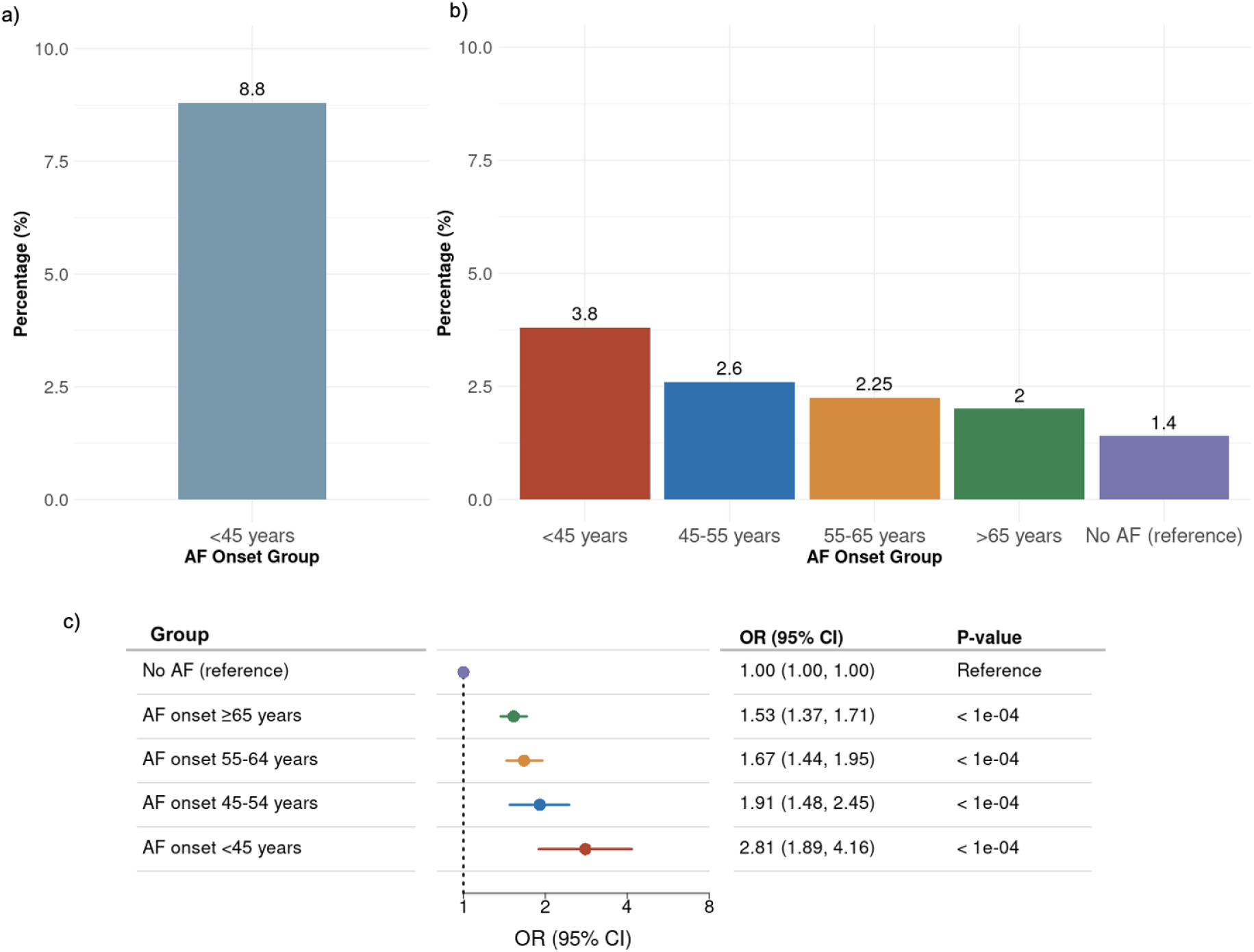
Percentage and odds ratio of participants carrying predicted Loss-of-Function (pLOF) variants in cardiomyopathy genes based on age at atrial fibrillation (AF) diagnosis. **Figure 1a:** Danish cohort data showing the percentage of carriers. **Figure 1b:** UK Biobank data stratified by age at AF onset. The highest prevalence of carriers is seen in individuals with AF onset before 45 years (red), while the lowest is seen in individuals with late-onset AF (green) or no diagnosis (purple). **Figure 1c:** Forest plot of UK Biobank data, displaying odds ratios (ORs), confidence intervals (CI) and p-values of carrying truncating variants, categorized by age at AF onset. Each group is compared to the reference group for statistical significance.

Of the 38 variants identified, 26 were classified as variants of unknown significance (VUS), while the remaining variants were categorized as pathogenic (five variants), pathogenic/likely pathogenic (six variants), and likely pathogenic (one variant), according to ClinVar(25).

The *TTN* gene was the most frequently affected, with 29 (69.0%) of the 42 individuals harboring a truncating variant (see **Supplementary Table S3**). *TTN* truncating variants accounted for the majority (n = 25, 65.8%) of all pLOF identified in the cohort. *LMNA* followed, with truncating variants observed in three individuals (7.9%), as shown in **Supplementary Table S3**. Furthermore, truncating variants were identified in two individuals for *RBM20*, *TPM1*, and *DSC2* (5.3% per gene), respectively, whereas *MYBPC3*, *PKP2*, *MYH7*, and *SCN5A* had variants in a single individual each (2.6% per gene). The distribution of disease-associated variants in this cohort revealed that 88.1% of all variants were associated with DCM genes (e.g., *TTN*, *LMNA*, *SCN5A, RBM20, MYH7 and TPM1*), 3.9% to HCM genes (*MYBPC3* and *MYH7*), and 7.9% to ARVC genes (*PKP2* and *DSC2*).

### Rare pLOF variants in the UK Biobank

A total of 634 individuals (2.17%) from the AF-diagnosed population in the UK Biobank were identified as carriers of rare truncating variants in established cardiomyopathy genes.

We observed an inverse dose-response relationship between the age of AF onset and the prevalence of rare pLOF variants, as shown in **Figure 1b**. The highest prevalence of carriers was found among individuals diagnosed with AF below the age 45, at 3.8%. In comparison, the prevalence was 2.6% for those with AF onset between 45 and 54 years, 2.2% for 55 to 64 years, 2.0% for those aged 65 and older, and 1.4% among individuals without an AF diagnosis.

Details on the truncating variant carrier counts are presented in **Supplementary Table S4**, where we report the variant count for AF patients with onset <45 years, the overall AF population, and the entire study population from UK Biobank. Additionally, percentages are reported based on the size of each group, providing a comparative distribution of truncating variants for the 29 selected genes across different cohorts.

Individuals with early-onset AF had a significantly higher OR for carrying a pLOF variant (**Figure 1c**). Compared with referents without AF, there was a 2.81-fold increase in OR for carrying a pLOF variant in individuals with AF onset before 45 years of age (95% CI 1.89-4.16; P <0.0001), representing the highest risk. This was followed by a 1.91-fold increase in individuals with onset between 45 and 54 years (95% CI 1.48-2.45; P<0.0001), a 1.67-fold increase with onset between 55 and 64 years (95% CI 1.44-1.95; P<0.0001), and a 1.53-fold increase with onset at or above 65 years (95% CI 1.37-1.71; P<0.0001).

Furthermore, we examined the distribution of variants across the 29 investigated cardiomyopathy genes in the UK Biobank (**Figure 2)**, and found that genes associated with DCM represented 57% of all variants present in the AF group in the UK Biobank, followed by 22% of variants related to ARVC and 21% to HCM. The majority of the truncating variants were found in *TTN*, representing 32.8% of all rare truncating variants identified in the AF population, followed by variants in *PKP2* (10.9%) and *ALPK3* (9.1%).

**Figure 2.**
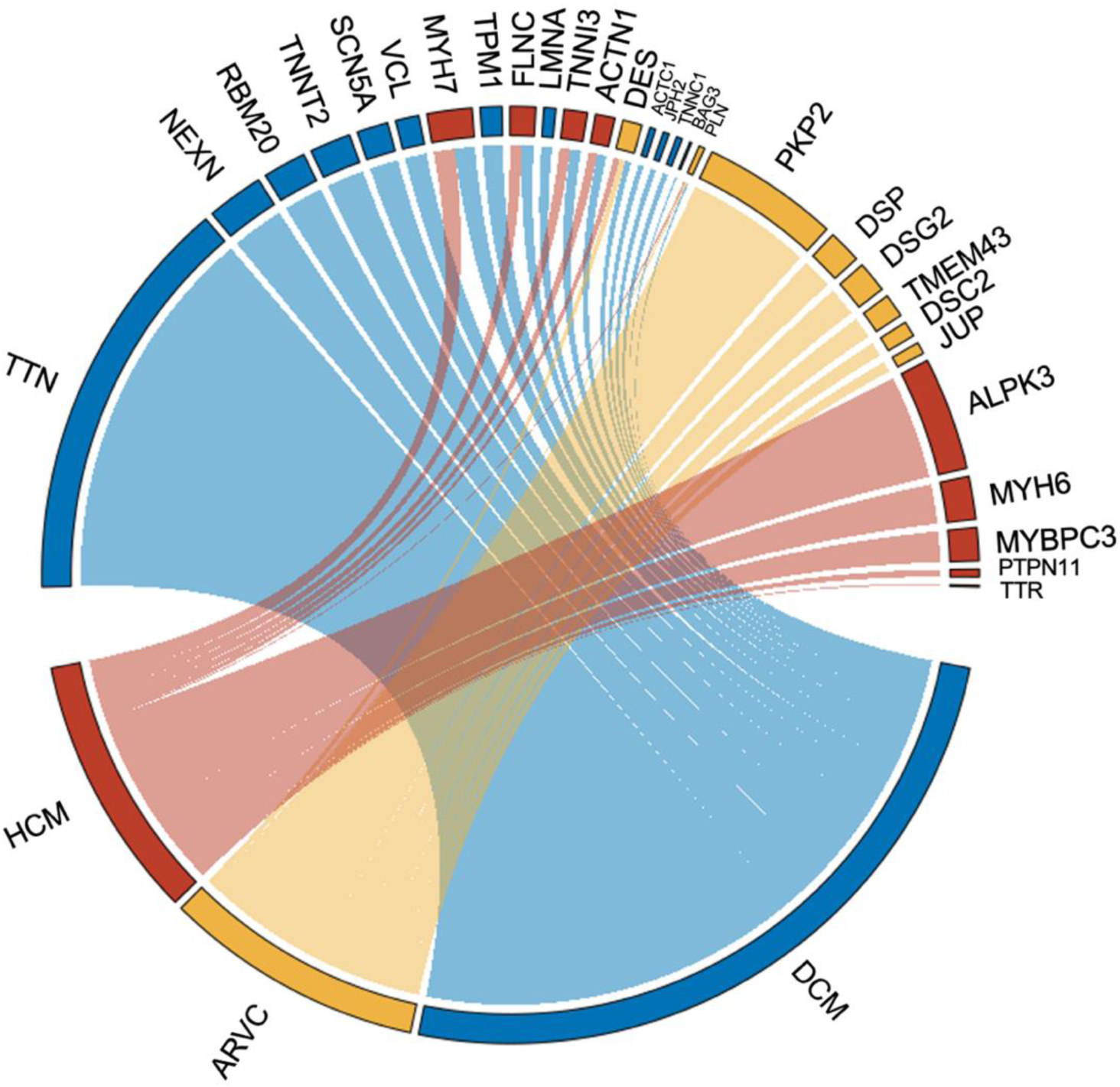
Distribution of rare predicted Loss-of-Function (pLOF) variants in individuals diagnosed with atrial fibrillation (AF) according to genes and their association with cardiomyopathy. Chord diagram illustrating the distribution of rare pLOF variants according to genes and corresponding cardiomyopathy subtypes. The blue segments represent variants in genes associated with dilated cardiomyopathy (DCM), the red segments correspond to variants in hypertrophic cardiomyopathy (HCM)-associated genes, and the yellow segments indicate variants in genes associated with arrhythmogenic right ventricular cardiomyopathy (ARVC).

### Missense Variants in the UK Biobank

We identified 389 individuals with AF (1.33%) from the AF-diagnosed population as carriers of rare missense variants (MTR < 5%) in the studied genes. The highest prevalence, 2.19%, was seen in individuals diagnosed below the age of 45 (**Figure 3a),** showing an inverse dose-response-like pattern. The prevalence rates for other age groups were 1.34% for 45 to 54 years, 1.31% for 55 to 64 years, 1.31% for those aged 65 and above, and 1.28% among individuals without an AF diagnosis. **Supplementary Table S5** provides the count of carriers among individuals with AF across age groups.

**Figure 3.**
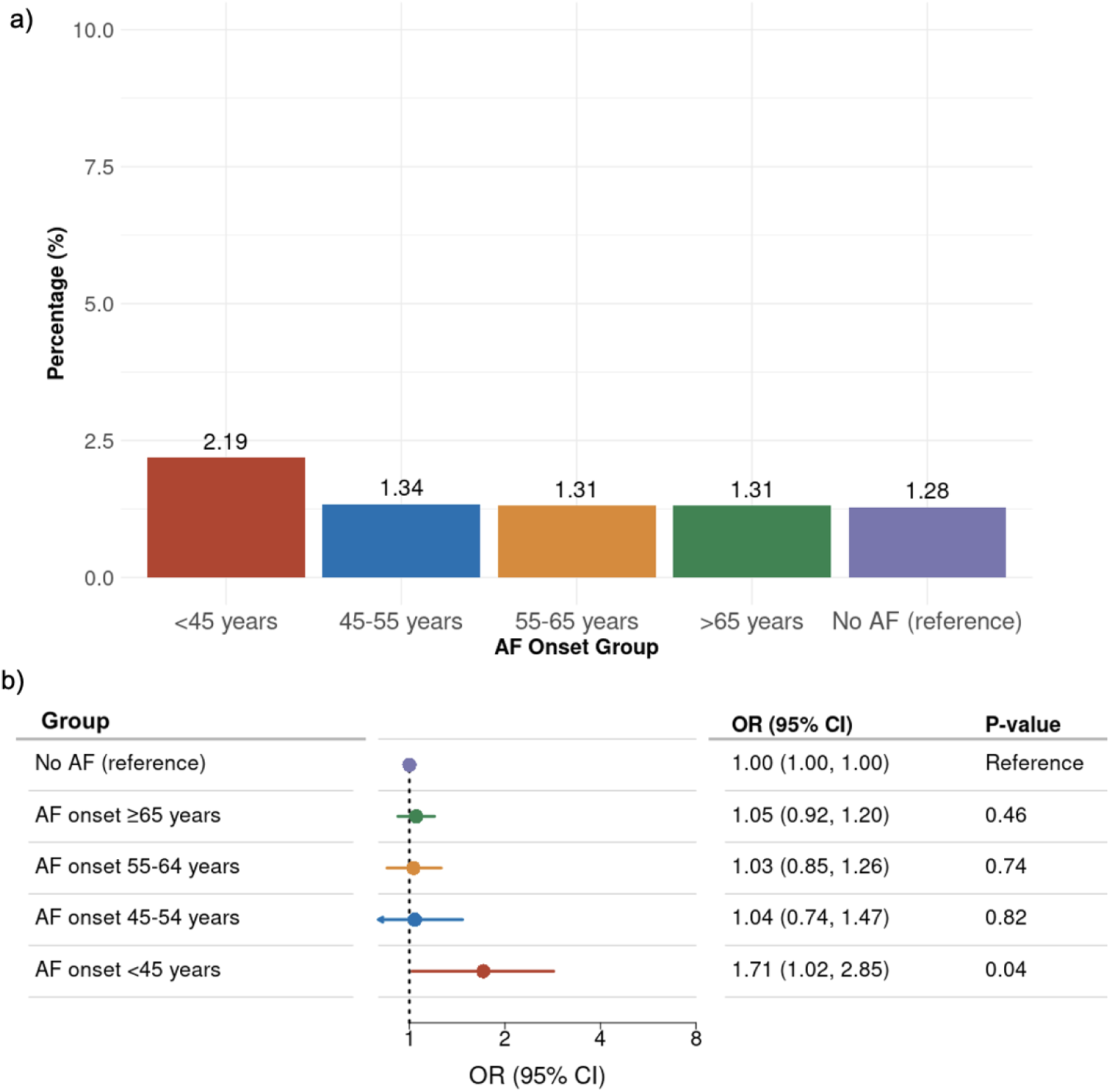
Percentage and odds ratio of participants carrying missense variants in cardiomyopathy genes based on age at atrial fibrillation (AF) diagnosis. **Figure 3a:** UK Biobank data showing rare missense variant carrier percentage, specifically for MTR below the 5^th^ percentile, stratified by age at AF onset. The highest prevalence of carriers is observed among individuals with AF onset before 45 years (red), while the lowest is seen in individuals with onset of AF at or after 65 years of age (green) or no diagnosis (purple). **Figure 3b:** Forest plot displaying odds ratios (ORs), confidence intervals (CI) and p-values for missense MTR below the 5^th^ percentile carriers in the UK Biobank.

**Figure 3b** illustrates the OR for carrying a missense variant among different age groups of AF onset. Carriers with AF onset before 45 years of age had a 1.71-fold increase in OR for carrying a variant (95% CI 1.02-2.85; P = 0.04), compared with referents without AF. No statistically significant associations were found for groups with later onset of AF.

Of the identified missense variants, 71.8% were in genes associated with HCM, 27.2% were in genes associated with DCM, and 1% were in genes associated with ARVC (Figure 4). The majority of these variants were identified in the *TTR* gene (54%), followed by *MYH7* (8.9%) and *FLNC* (7.4%).

**Figure 4.**
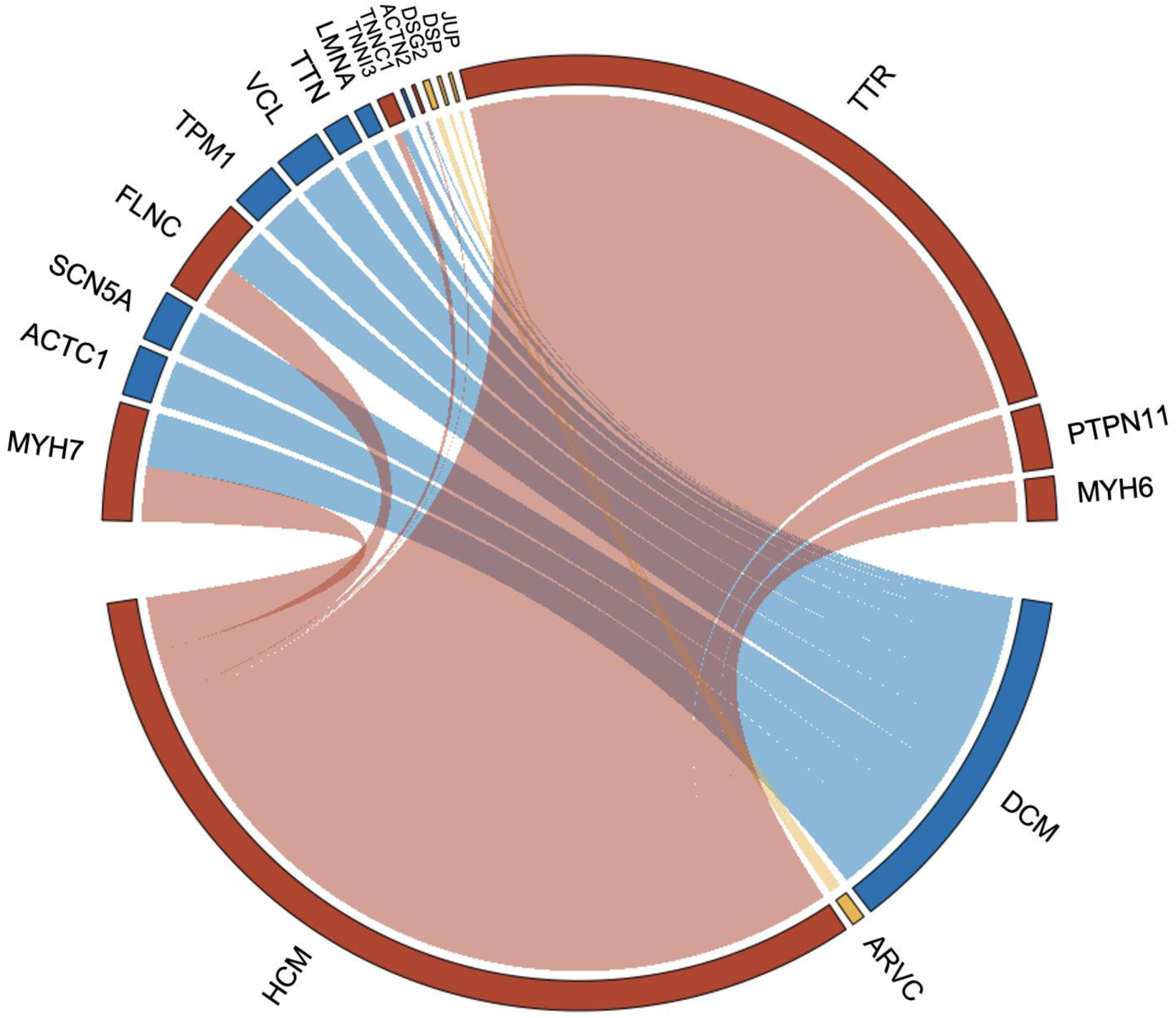
Distribution of rare missense variants in individuals diagnosed with atrial fibrillation (AF) according to genes and their association with different cardiomyopathies. The blue segments represent variants in genes associated with dilated cardiomyopathy (DCM), the red segments correspond to variants in hypertrophic cardiomyopathy (HCM)-associated genes, and the yellow segments indicate variants in genes associated with arrhythmogenic right ventricular cardiomyopathy (ARVC).

### Aggregated prevalence of genetic variants in the UK Biobank

Integrating both truncating and missense variants indicated a larger aggregated percentage of carriers while following the same pattern. A total of 982 individuals (3.37%) from the AF population carried genetic variants (see **Table S6 in the Supplementary**).

**Figure 5a** shows a consistent pattern, with the highest prevalence seen in early-onset AF individuals at 5.99%. In comparison, the combined prevalence was 3.86% in individuals with AF onset between 45 and 54 years, 3.52% in the 55 to 64 years group, 3.30% in those aged 65 and above, and 2.66% among individuals without an AF diagnosis.

**Figure 5.**
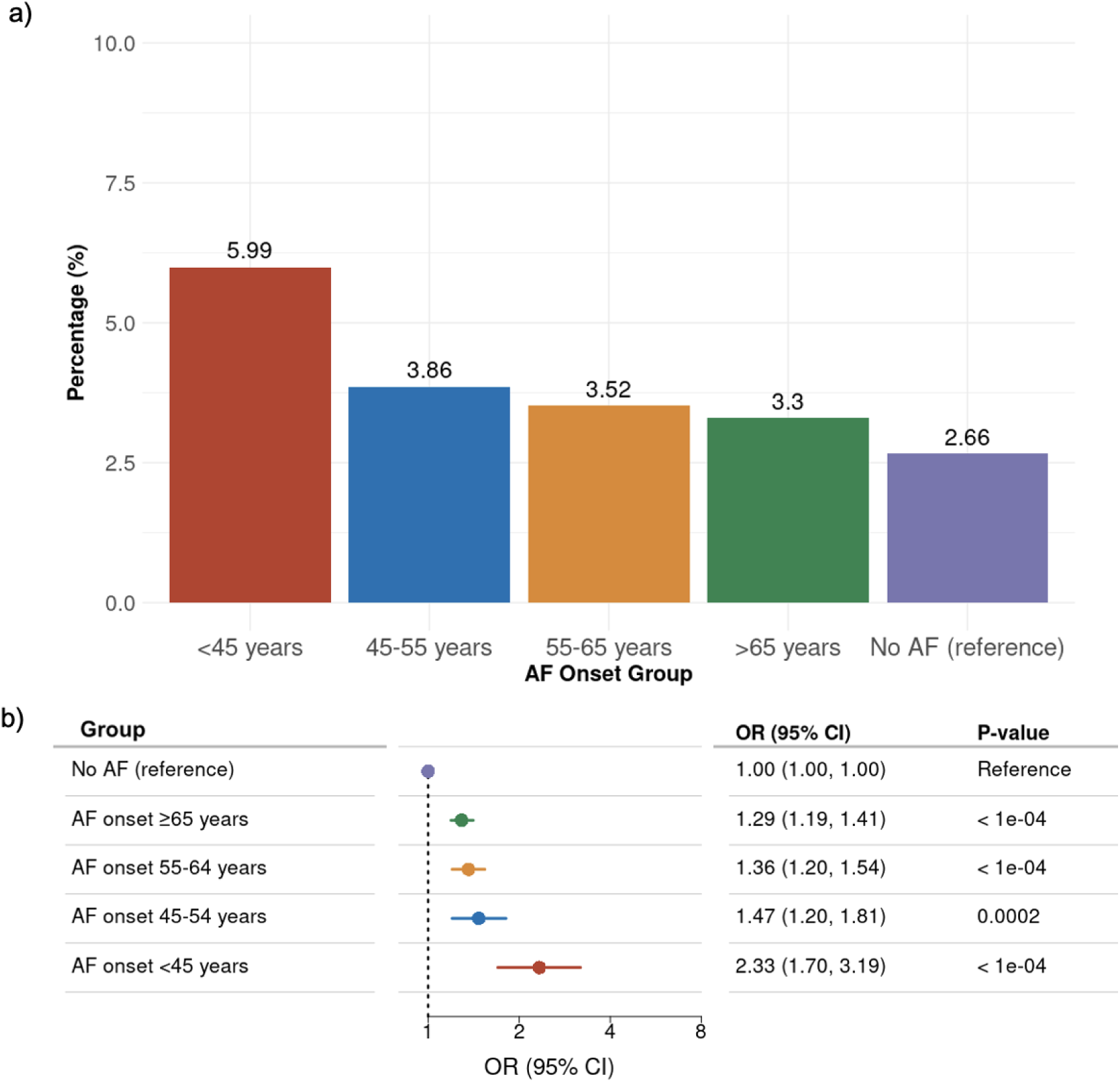
Prevalence and odd ratios of rare missense and predicted Loss-of-Function variants in cardiomyopathy genes according to age at atrial fibrillation diagnosis. **Figure 5a:** UK Biobank data on rare truncating and missense variant carrier percentages stratified by age of onset. **Figure 5b:** Forest plot displaying ORs, CI and p-values for the carriers of truncating and missense variants, comparing AF onset groups to the reference population in the UK Biobank.

**Figure 5b** shows the aggregated OR of carrying either rare truncating or missense variants within the UK Biobank population across different age of AF onset groups.

## Discussion

The present study combined data from a cohort of early-onset AF patients with large-scale clinical and genetic information from the UK Biobank, to elucidate the prevalence of genetic variants in known cardiomyopathy genes in patients with AF. We observed that early-onset AF was significantly associated with a greater prevalence of rare genetic variants in the 29 genes associated with either DCM, HCM or ARVC.

Among the Danish individuals with early onset AF, more than one in twelve (8.8%) carried a rare truncating variant. The majority of these variants were in DCM-associated genes, with *TTN* variants accounting for 65.8% of all truncating variants. Nearly two-thirds of those identified were variants of unknown significance. *TTN* encodes titin, a key structural protein in the sarcomere, and truncating variants in this gene are well-established contributors to the development of DCM(26) and AF.

Following *TTN*, *LMNA* was the second gene with the most variants in this cohort, representing 7.9% of the variants found. Variants in *LMNA*, which encodes lamin A/C, are commonly associated with early-onset AF and are linked to severe complications such as DCM and sudden cardiac death(27–29). Additionally, we identified a single nonsense variant in *SCN5A*. While this sodium channel is primarily susceptible to missense variants that are associated with atrioventricular block and DCM(29,30), the nonsense variant we identified has been classified as pathogenic as it alters intracellular sodium and calcium homeostasis(26). As previously reported(4), we also observed two pLoF variants in *RBM20*, a regulator of splicing for various cardiac genes, including *TTN* and *TPM1*(4). *TPM1*, encoding the actin-binding tropomyosin 1 protein, exhibited two variants in this Danish cohort. Additionally, one individual carried a truncating *MYBPC3* variant, and another individual carried a nonsense variant in *MYH7*. Notably, *MYBPC3* and *MYH7* are known to be involved in 70% of sarcomere gene variant-positive HCM(31).

Furthermore, we identified three variants in two desmosomal genes associated with ARVC, one in *PKP2* and two in *DSC2*. *PKP2* encodes plakophilin 2, a desmosomal protein crucial in ARVC, where it disrupts intracellular calcium handling and predisposes patients to ventricular arrhythmias(26). *DSC2*, a known ARVC disease susceptibility gene, is essential for cardiac desmosome formation, early cardiac morphogenesis, and normal cardiac function(32).

Analyses in the UK Biobank offered insights into the prevalence and distribution of genetic variants across different age groups. Specifically, we found an OR of 2.8 for carrying truncating variants in the individuals with early-onset AF compared to the reference group. Previous studies have described a similar pattern(6). In our study, we focused on constitutive exons with a TTN PSI >90(33) and selected variants with a MAF of <1%(34). This approach allows us to focus on rarer, potentially more pathogenic variants that are likely to have a greater impact on disease development. The majority of these truncating variants were found in key genes associated with DCM (e.g. *TTN* and *RBM20*), HCM (e.g. *ALPK3, MYH6* and *MYBPC3*), and ARVC (e.g. *PKP2, DSP* and *DSG2*). In this cohort, variants in genes associated with DCM accounted for 60% of carriers, driven largely by the high prevalence of *TTN* truncating variants(35–37). Other prominent contributors included well-established DCM-related genes such as *NEXN*(38), *MYH7*(6,29), *RBM20*(4,6,29)*, TNNT2*(29) and *SCN5A*(26,29,30).

In contrast to our findings from the Danish cohort, *LMNA* variants were relatively rare in the UK Biobank cohort. One possible explanation is that *LMNA* variants typically lead to early-onset DCM(27), often manifesting before the age range of individuals included in the UK Biobank, which may have resulted in the underrepresentation of these variants.

Beyond DCM, 16% of the truncating variant carriers harbored HCM-associated variants in genes including *ALPK3*(39)*, MYH7*(40), *MYH6*(18) and *MYBPC3* (41). The majority of variants in HCM genes were in *MYH7,* a sarcomeric gene encoding the β-myosin heavy chain, known to be implicated in both DCM and HCM depending on the penetrance(40,41), accounted for a significant proportion of carriers.

In total, 24% of UK Biobank participants diagnosed with AF and carriers of genetic variants had truncating variants in genes previously linked to ARVC, with *PKP2* being the most frequent. Additional variants were found in other ARVC-associated genes such as *DSP*, *DSG2*, and *TMEM43*(26), all of which encode desmosomal proteins. Defects in the structure of desmosomes result in cardiac myocyte detachment and cell death, leading to fibrofatty tissue infiltration and electrical instability(45).

To elucidate a more detailed genetic landscape of AF across age groups, we also examined rare missense variants, applying an MTR score to focus on highly conserved regions. While the overall prevalence of missense variants showed a similar trend to that of truncating variants, their impact was less pronounced, with early-onset AF patients having a 1.7-fold increased OR of carrying these variants compared to the reference group. Missense variants were predominantly found in genes associated with HCM, accounting for 71% of carriers. *MYH7* had the highest prevalence of missense variants in this population, emphasizing the relevance of both missense and truncating variants in *MYH*7.

A higher prevalence of rare missense variants was observed in the *TTR* gene compared to truncating variants. *TTR* encodes the protein transthyretin, and mutations in this gene have been associated with cardiac amyloidosis (ATTR-CA) and HCM(46). Another significant gene in the missense carrier group was *FLNC*. A previous study showed that *FLNC* missense variants were predominantly associated with HCM, while truncating variants were rather strongly associated with DCM(47).

The analysis of missense variants across cardiomyopathies revealed notable differences compared to truncating variants. Truncating variants were most frequently found in genes associated with DCM, whereas HCM and ARVC genes had a similar prevalence of missense and truncating variants. In contrast, disease-associated missense variants were most commonly found in HCM genes, followed by DCM genes, while missense variants in ARVC-associated genes were rare. These findings hint at distinct genetic mechanisms underlying each variant type and their respective roles in specific cardiomyopathies.

In summary, our findings indicate that individuals with very early onset of AF have a high prevalence of potentially pathogenic variants in cardiomyopathy-associated genes. Thus, these individuals may benefit from more thorough clinical monitoring to aid in identifying individuals at an elevated risk of developing cardiomyopathy and subsequent HF. While systematic genetic screening for rare variants is unlikely to be implemented in the near future, incorporating genetic data into clinical practice could improve cardiomyopathy risk stratification in individuals with AF.

## Limitations

The findings from this study should be interpreted with the following limitations in mind. First, the Danish cohort only included individuals with very early-onset AF, which is not representative of the typical patient with AF. This selection may partly explain the higher prevalence of truncating variants observed in the Danish cohort compared to individuals with early-onset AF in the UK Biobank. Additionally, a healthy-volunteer bias in the UK Biobank may lead to an underrepresentation of individuals with pathogenic genetic variants, potentially limiting the generalizability of these findings. Furthermore, targeted sequencing of the Danish cohort did not include *ALPK3*, *ACTC1*, *TNNC1*, and *PLN*, which may affect the comprehensiveness of our variant analysis.

Moreover, not all variants may be equally pathogenic or penetrant. As participants had to survive until inclusion in the respective cohorts, this could introduce immortal time bias and result in the underrepresentation of certain variants compared to others. Finally, both cohorts were of white European ancestry, and it cannot be assumed that the findings can be generalized to other population groups.

## Conclusion

By using two different AF cohorts, we found that a significant number of individuals with early onset of AF carried rare pathogenic variants in hallmark cardiomyopathy genes. Prevalence of truncating variants ranged from one in twelve, to one in fifteen of individuals with AF onset below age 45. These findings support that screening and clinical surveillance for cardiomyopathy might be relevant for individuals with early-onset AF.

## Data Availability

Due to privacy concerns, data underlying the Danish AF cohort cannot be shared. Data from the UK Biobank is available for bona-fide researchers through application to https://www.ukbiobank.ac.uk/enable-your-research/apply-for-access

## Acknowledgments

The UK Biobank was accessed under application number 43247. The authors would like to thank all participants from both cohorts for their participation. Our study integrates publicly available data based on the gnomAD and TOPMed project, and we would like to acknowledge the investigators and participants of these studies.

## Non-Standard Abbreviations and Acronyms

AF: Atrial Fibrillation
DCM: Dilated Cardiomyopathy
HCM: Hypertrophic Cardiomyopathy
ARVC: Arrhythmogenic Right Ventricular Cardiomyopathy
HF: Heart failure
pLoF: Predicted Loss-of-Function
MTR: Missense Tolerance Ratio
OR: Odds Ratio
CI: Confidence Interval
MAF: Minor Allele Frequency
VUS: Variant of Uncertain Significance
PSI: Percent Spliced In
ACMG: American College of Medical Genetics and Genomics
ACC/AHA: American College of Cardiology and the American Heart Association

